# New evidence that *Callithrix* spp. is assuming an important role in the epidemiology of yellow fever virus

**DOI:** 10.1101/2025.09.07.25335112

**Authors:** Márcio Junio Lima Siconelli, Jéssica Caroline de Almeida Dias, Eduardo Ferreira Machado, Mariana Sequetin Cunha, Natália Coelho Couto de Azevedo Fernandes, Juliana Mariotti Guerra, Luana Bonon, Alline Borges Salomão, Karin Werther, Karina Paes Bürger, Adolorata Aparecida Bianco Carvalho, Daniel Marques, Benedito Antonio Lopes da Fonseca

## Abstract

In 2023, a new yellow fever virus (YFV) lineage was introduced in São Paulo State, Brazil. From July 2024 to June 2025, nine *Callithrix penicillata* tested positive, showing low YFV viral load but characteristic organ lesions, suggesting this genus may replace *Alouatta* sp. as sentinel to detect YFV circulation.

---

Between 2016-2018 Brazil experienced the worst outbreak of wild yellow fever in the last 70 years. The southeast region was responsible for most human and non-human primate (NHP) cases (1, 2). *Orthoflavivirus flavi* [yellow fever virus (YFV)] is an arbovirus from Flaviviridae family transmitted by *Aedes* and *Haemagogus* or *Sabethes* mosquitoes in the urban and sylvatic cycles, respectively.

NHPs are key sentinel animals for early YFV detection and a core component of Brazilian surveillance program to prevent human cases (3). However, YFV pathogenicity varies by NHP genus and species and its importance across Brazil’s diverse NHPs remains poorly understood (4). Notably, primates from *Alouatta* genus (howler monkeys) are the most susceptible to infection (5, 6, 7), developing clinical and pathological manifestations, similar to those of humans (4, 8, 9). Infected individuals typically exhibit high viral loads and pronounced liver damage, including areas of necrosis and apoptosis, Councilman–Rocha Lima (CRL) bodies, steatosis, and inflammatory infiltrates (10, 11).

In the last outbreak, the largest number of NHPs studied belonged to the *Callithrix* genus, which, revealed two distinct infection patterns, one with a high viral load and typical lesions and a second, seen more frequently, with low viral loads, absence of clinical disease, and either absent or minimal hepatic lesions (4, 12).

In 2023, a new YFV lineage was introduced in São Paulo State, initially detected in the eastern region (13) and then spread to other regions (northeast and then northwest). The first confirmed NHP deaths attributed to YFV in the northwest region (Ribeirão Preto city) occurred in late December 2024. Over the following months, the outbreak intensified, with more confirmed NHPs infections in surrounding municipalities (figure 1). During this period one human case was recorded in the region.

**Figure 1.**
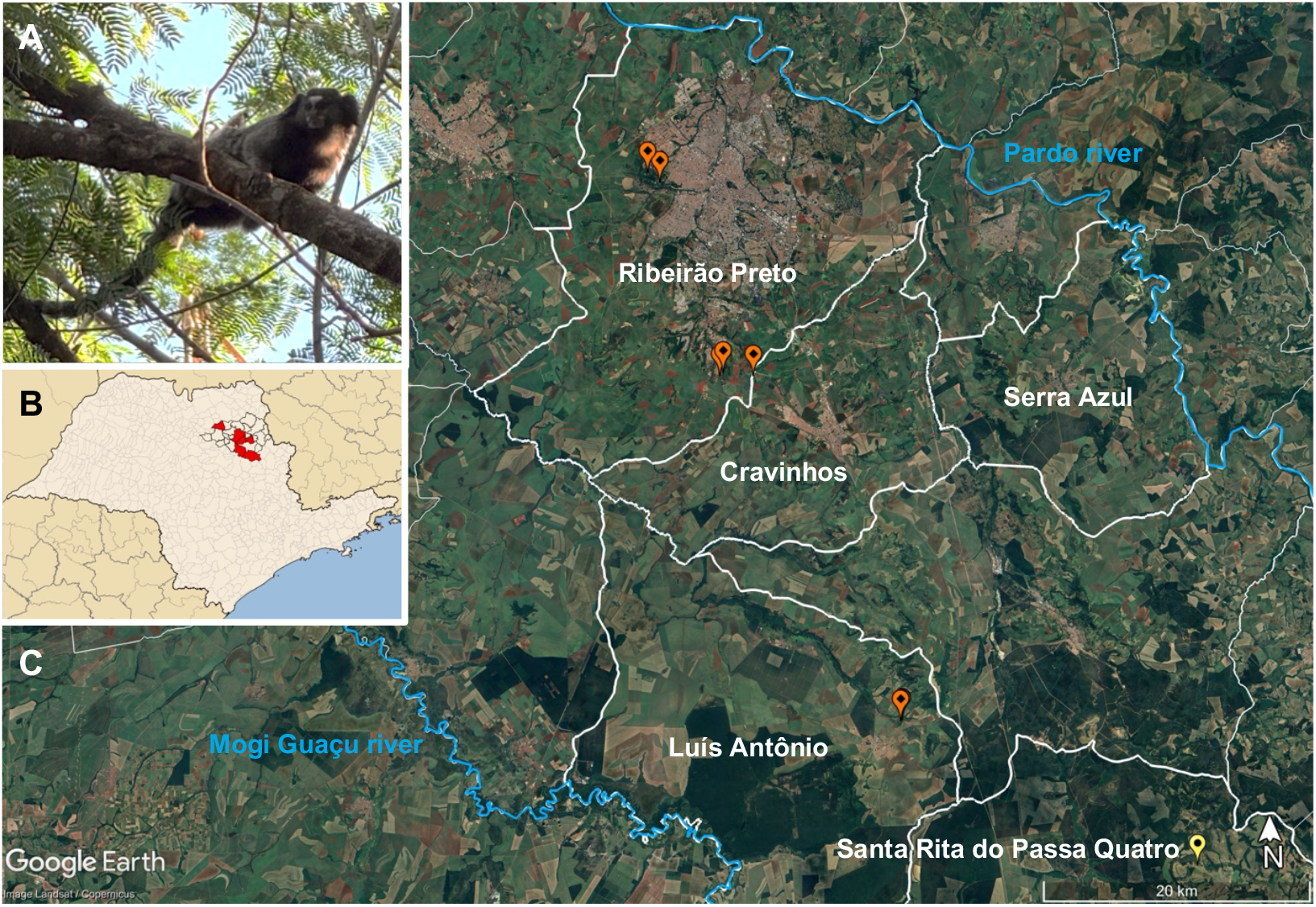
**A)** free-living animal *C. penicillata* species recorded during fieldwork. **B)** São Paulo State map with dark lines highlighting the municipalities of the regional Epidemiological Surveillance Group and in red the affected areas by YFV. **C)** Partial aerial view of the Ribeirão Preto region. Stronger white outlines represent the borders of the municipalities where YFV was detected between July 2024 and June 2025. Orange dots: locations where positive NHPs of the species *Callithrix penicillata* were found; Yellow dot: confirmed human yellow fever case. Sources: Google Earth, 2025 and São Paulo State municipalities map by Raphael Lorenzeto de Abreu, https://commons.wikimedia.org/w/index.php?curid=1119354, modified by the author.

## The Study

Between July 2024 and June 2025, the regional Epidemiological Surveillance Group reported 233 NHP deaths, of which 79% (184/233) were from *Callithrix penicillata* species, 18% (42/233) *Alouatta caraya*, 2.15% (05/233) *Sapajus nigritus* and 0.85% (02/233) *Callicebus nigrifrons*. Approximately 66% (154/233) were necropsied and biological samples were collected for diagnosis. Of these 88.9% (137/154) were Callitrichids.

Yellow fever was diagnosed in 54 NHPs, 22 by laboratory tests and 32 by epidemiological criteria. Among those confirmed by laboratory tests, 72.7% (16) were adults and 27.3% (6) juveniles. Females represented 45.45% (10/22). Positive NHPs belonged to *A. caraya* (50%; 11/22), followed by *C. penicillata* (40.9%; 9/22) and *C. nigrifrons* (9.1%; 2/22). As expected, samples from genera *Alouatta* and *Callicebus* had low cycle threshold (Ct) values and typical organ lesions including extensive hepatocellular necrosis, CRL bodies, hepatic steatosis, varying degrees of inflammatory infiltrate, and detectable YFV antigens by immunohistochemistry (IHC). The positive marmosets exhibited a consistent pattern: low Ct values, ranging from 11.14 to 20.08, typical lesions and detectable YFV antigens. One animal, stored frozen for a month before necropsy, showed a higher Ct value (28.9). All *C. penicillata* individuals were positive in IHC (figure 2). Two animals (22%; 2/9) showed advanced autolysis, which precluded evaluation of histopathologic features except for the presence of CRL bodies in one. Histopathologic evaluation in the remaining seven animals showed hepatic necrosis in all evaluated animals, with panlobular distribution ranging from multifocal (42%; 3/7) to widespread involvement (57%; 4/7). Necrosis severity ranged from moderate (29%; 2/7) to marked (71%; 5/7). CRL bodies were identified in all evaluated animals (100%; 7/7). Inflammatory infiltrates, present in 43% (3/7), consisted of mixed cellular contents in one case (33.3%; 1/3) and mononuclear cells in two cases (66.7%; 2/3), with multifocal distribution and moderate severity in two cases (66.7%; 2/3), and discrete in one (33.3%; 1/3). Hemorrhage was identified in 57% (4/7), characterized by diffuse distribution in most [75% (3/4)] and moderate to marked severity [75% (3/4) moderate; 25% (1/4) marked]. Steatosis was present in six cases [86% (6/7)], with both macrovesicular and microvesicular patterns, always panlobular, and moderate severity in 34% (2/6) or marked in 66% (4/6).

**Figure 2.**
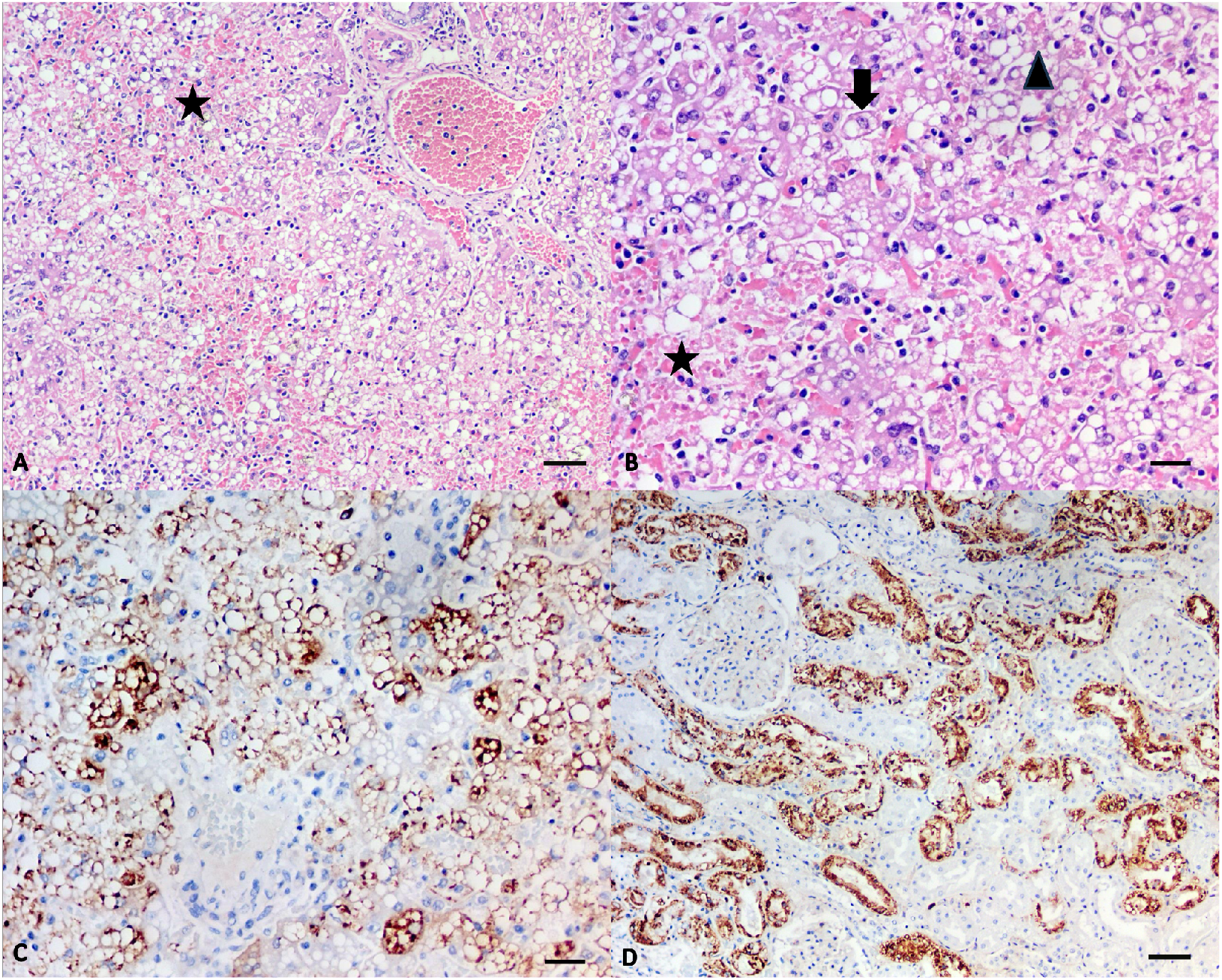
**A–D)** *Callithrix penicillata* with severe hepatic lesions due to yellow fever. **A)** H&E, 10X, liver. Acute and severe hepatic damage characterized by diffuse macrovesicular steatosis and focal hemorrhage (star). Scale bar = 40 µm. **B)** H&E, 20X, liver. Acute and severe hepatic damage with diffuse macrovesicular steatosis (black arrow), focal hemorrhage (star), and a Councilman–Rocha Lima body (alonged arrowhead). Scale bar = 20 µm. **C)** IHC, 20X, liver. Hepatocytes positive for yellow fever virus antigen (brown staining). Scale bar = 20 µm. **D)** IHC, 10X, kidney. Renal tubular epithelial cells positive for yellow fever virus antigen (brown staining). Scale bar = 40 µm.

During the 2017-2018 outbreak seven *Callithrix* spp. were positive to YFV but none had lesions or antigen detected (14). Of these, five were from Ribeirão Preto and the surrounding area. In 2020, an urban *C. penicillata* from the Brazilian midwestern region presented the same pattern, positive to YFV by RT-qPCR with no liver lesions or antigen detection (15).

The histopathological findings in association with IHC and molecular results of this study demonstrate a consistent pattern of severe acute hepatitis in neotropical primates infected with YFV during the 2024/2025 outbreak in São Paulo State. All 22 confirmed cases revealed characteristic hepatic lesions, with hepatocellular necrosis in all evaluated specimens, predominantly showing diffuse and multifocal to coalescing patterns. These findings corroborate with previous reports describing panlobular necrosis as a hallmark of fatal YFV infection in primates. CRL bodies, characteristic for YFV lesions, were identified in around 80% (18/22) of cases, with significantly higher frequency in *A. caraya, C. penicillata* and *C. nigrifrons*. Notably, even in specimens with marked autolysis these inclusion bodies remained detectable in three NHPs, reinforcing their diagnostic value even under suboptimal tissue preservation conditions. Inflammatory infiltrates were predominantly mixed (polymorphonuclear and mononuclear) or mononuclear, with panlobular distribution followed by periportal/portal. Macrovesicular, microvesicular or mixed steatosis with diffuse or multifocal distribution and consistently panlobular zonation was verified. Severity was marked to moderate in more than 72% (13/18) of affected animals.

The high prevalence and severity of lesions combined with limited inflammatory response, despite extensive necrosis and steatosis, may indicate a YFV-induced metabolic dysfunction and support direct viral injury as the main mechanism of liver damage. These unprecedented findings need further investigation as they point at a possible association with the new lineage of YFV circulating in southern Brazil.

A particularly relevant finding that supported this hypothesis was the IHC confirmation of active YFV infection in 100% of cases, including all analyzed *C. penicillata* specimens. This contrasts with prior observations (e.g., 2017–2018), where *Callithrix* spp. often exhibited lower viral loads and milder hepatic lesions (4, 14, 15). In the NHPs evaluated here, marmosets developed severe liver damage comparable to howler monkeys, with extensive necrosis and unequivocal IHC positivity. The severe lesions observed in *Callithrix* underscore its importance in a surveillance based on multiple species, particularly in scenarios where Alouatta spp. populations have been significantly reduced by the latest YFV outbreak.

## Conclusions

Our results confirm the classic histopathological profile of YFV infection in neotropical primates, with striking severity in *C. penicillata* during this outbreak. The combination of extensive hepatocellular necrosis, CRL bodies, marked steatosis, and 100% IHC positivity provides robust evidence of YFV pathogenicity across multiple primate species and reinforce the importance of continuous surveillance in NHPs, placing the genus *Callithrix* under the public health spotlight now and in the futures epidemics. Its ability to adapt to urbanized environments, its proximity to humans and the high population of *Aedes aegypti* mosquitos constantly raises the fear of YFV reurbanization. These findings are important since they may represent the need for the establishment of a new surveillance system based on the *Callithrix species*.

Therefore, local zoonotic surveillance services in high-risk areas must be strengthened to detect early YFV circulation in the face of NHP deaths, especially in view of a possible change in the epidemiological pattern of amplifying hosts, with marmosets assuming a role previously occupied by howler monkeys.

## Data Availability

All data produced in the present work are contained in the manuscript

## Ethics

Animal samples were collected by the zoonotic surveillance unit of each municipality as part of national surveillance program of yellow fever and sent to Molecular Virology Laboratory of Ribeirão Preto Medical School – University of São Paulo for initial detection of YFV and to Adolfo Lutz Institute to confirm the diagnosis. As Yellow Fever is a disease with a high impact to the public health system, every detection of YFV needs to be confirmed by a national reference laboratory that, in São Paulo state, is the Adolfo Lutz Institute. All animals were found dead naturally. This study does not require approval by Ethics Committees according to Brazilian normative resolution n^∘^ 30, of February 2, 2016 of the National Animal Experimentation Control Council - CONCEA.

## Acknowledgments

We would like to thank all the municipal zoonotic surveillance services of XXIV Health Region of São Paulo state, particularly the municipalities of Ribeirão Preto, Cravinhos, and Luís Antônio, for their prompt response and for providing the necessary samples for official diagnostic confirmation.

## Author’s contributions

**MJLS**: Conceptualization, Methodology, Investigation, Data Curation, Writing - Original Draft, Writing - Review & Editing, Project administration, Funding acquisition; **JCAD**: Investigation and Samples Processing; **EFM**: Data Curation, Writing - Original Draft, Writing - Review & Editing; **MSC, NCCAF** and **JMG**: Investigation, Sample Processing and Writing - Review & Editing; **LB** and **ABS:** investigation and sample collection; **KW, KPB, AABC** and **DM**: Writing - Review & Editing **BALF**: Conceptualization, Methodology, Resources, Writing - Review & Editing, Supervision, Project administration, Funding acquisition. All authors read, reviewed and approved the final version of the manuscript.

